# Transseptal or Transapical Transcatheter Valve-in-Valve Implantation Versus Redo Surgical Replacement for Degenerated Mitral Bioprostheses

**DOI:** 10.1101/2025.04.08.25325495

**Authors:** Pedro Felipe Gomes Nicz, Mauricio Felippi de Sá Marchi, Henrique Barbosa Ribeiro, Fernando Chiodini Machado, Roney Orismar Sampaio, José Honório de Almeida Palma da Fonseca, José Airton de Arruda, Carlos Henrique Eiras Falcão, Maurício Lopes Prudente, Vitor de Andrade Vahle, Arnaldo A Okino, Marco Antônio Praça de Oliveira, Maurílio Onofre Deininger, Antenor Lages Fortes Portela, Salvador André Bavaresco Cristovão, Rodrigo de Castro Bernardes, Breno de Alencar Araripe Falcão, Estêvão Carvalho de Campos Martins, Luiz Eduardo Koenig de São Thiago, Adriano Dias Dourado Oliveira, Fabio Solano de Freitas Souza, Eduardo França Pessoa de Melo, Antônio Carlos Botelho da Silva, João Eduardo Tinoco de Paula, Evandro Martins Filho, Ari Mandil, Mateus S Viana, Rogério Sarmento-Leite, Wilton Francisco Gomes, Cristiano Guedes Bezerra, Antonio José Neri de Souza, Valter Correa Lima, Moysés de Oliveira Lima-Filho, Roger Renault Godinho, Gabriela Liberato de Souza, David Le Bihan, Wilson Mathias, Pablo Pomerantzeff, Fábio Biscegli Jatene, Roberto Kalil-Filho, Flávio Tarasoutchi, Azeem Latib, Alexandre Abizaid, Fábio Sandoli de Brito

## Abstract

**Background:** Bioprosthetic mitral valve degeneration can be problematic as redo surgical mitral valve replacement (Redo-SMVR) is associated with a high morbidity and mortality. Valve-in-valve transcatheter mitral valve replacement (TMVR) has emerged as an alternative for high-risk patients, but comparative data is lacking.

**Objectives:** We aimed to evaluate the outcomes of Redo-SMVR vs. TMVR, using either transapical (TA) or transseptal (TS) approach.

**Methods and Results:** A total of 415 patients (69% rheumatic) were included between January 2014 to May 2023 (Redo-SMVR=239; TA-TMVR=84; TS-TMVR=92). Patients in the Redo-SMVR group were younger (51.7±12.3, 64.3±10.5, 73.7±10.9, respectively for Redo, TA and TS; p<0.001) with a lower median STS score (2.9 [1.7 – 5.1], 6.6 [3.9–10.5], 6.5 [4.2–10.0]; p<0.001). 30-day mortality was significantly higher for both Redo-SMVR and TA-TMVR vs. TS-TMVR (14.2%, 15.5%, 4.3%; p=0.030) and procedural success was lower (52.5%, 64.9%, 88%; p<0.001). There was no difference in terms of 1-year mortality rate across the groups (Log-rank p=0.222). Reduced preprocedural left ventricular ejection fraction, higher right ventricular systolic pressure (RVSP), and the presence of coronary artery disease were independently associated with 30-day mortality, while transseptal approach was protective. At 1-year follow-up, the predictors of mortality remained consistent, with significant tricuspid regurgitation replacing RVSP (all with p<0.05).

**Conclusions:** In high-risk patients with dysfunctional mitral bioprosthesis and most with rheumatic etiology, TS-TMVR was associated with higher procedural success and better short-term outcomes, including all-cause mortality, despite higher risk characteristics. In the mid-term follow-up, there were no significant differences in mortality across the groups.

## INTRODUCTION

Mitral valve disease is a common valvular heart disease,^1^ with a high prevalence of rheumatic heart disease in some developing countries, contributing to its earlier presentation in these regions.^2^ Surgical mitral valve repair or replacement (SMVR) is the treatment of choice for the majority of such patients, with a growing use of bioprosthetic valves in the recent decades, due to a reduced need for anticoagulation and lower thrombotic complications, as compared with mechanical prostheses.^3-5^ Hence, an increasing number of patients develop bioprosthetic mitral valve degeneration,^6^ leading to reintervention rates of up to 35% at 10-years follow-up.^7^ Redo-SMVR has been the treatment of choice for mitral bioprosthetic degeneration, however it is associated with significant morbidity and periprocedural mortality, especially in older patients and with concurrent comorbidities.

Therefore, valve-in-valve transcatheter mitral valve replacement (TMVR) emerged as a less invasive procedure, performed through either the transapical (TA) or transseptal (TS) approaches.^8,9^ Two prior large studies, one from the VIVID Registry^10^ and another from the TVT Registry^11^, have determined the overall safety and efficacy of the TMVR, and together with other smaller studies,^12-14^ have permitted this indication by major regulatory agencies as an alternative to Redo-SMVR for high-risk patients. Additionally, retrospective registries have compared Redo-SMVR vs. TMVR for mitral bioprosthetic valve failure, irrespective of the approach used (TA or TS), but still with limited number of patients and events, and no direct comparison of the approach used (TA vs. TS).^15-20^ The present study aimed to investigate the 30-day and 1-year clinical and echocardiographic outcomes of Redo-SMVR, TA-TMVR and TS-TMVR in a real-world patient level registry.

## METHODS

### Study design and patient population

This is a multicentre observational study that enrolled consecutive patients presenting with mitral bioprosthetic valve failure who underwent TS-TMVR, TA-TMVR or Redo-SMVR. TS-TMVR was performed in all 31 Brazilian centers from the National Registry run by the Brazilian Society of Interventional Cardiology. TA-TMVR, in addition to Redo-SMVR were exclusively performed at one of the 31 institutions, the Heart Institute – University of Sao Paulo Medical School (InCor).^14^

All patients had severely degenerated mitral bioprostheses, and the procedural indication was made according to the local Heart Team decision, taking into consideration clinical and anatomical features. Patients who presented with any contraindication to the transcatheter approach, such as active endocarditis and valve thrombosis, were excluded. The exclusion criteria also encompassed patients with incomplete medical records as well as combined disorders requiring intervention, except for tricuspid regurgitation.

Transcatheter procedures started in May 2015, when the first TA-TMVR procedure was performed, while the first TS-TMVR was done in June 2016. Redo-SMVR cases were collected since 2014. The number of procedures fluctuated throughout the years, while TA-TMVR procedure fell and TS-TMVR rose more recently (**Figure 1**).

**Figure 1.**
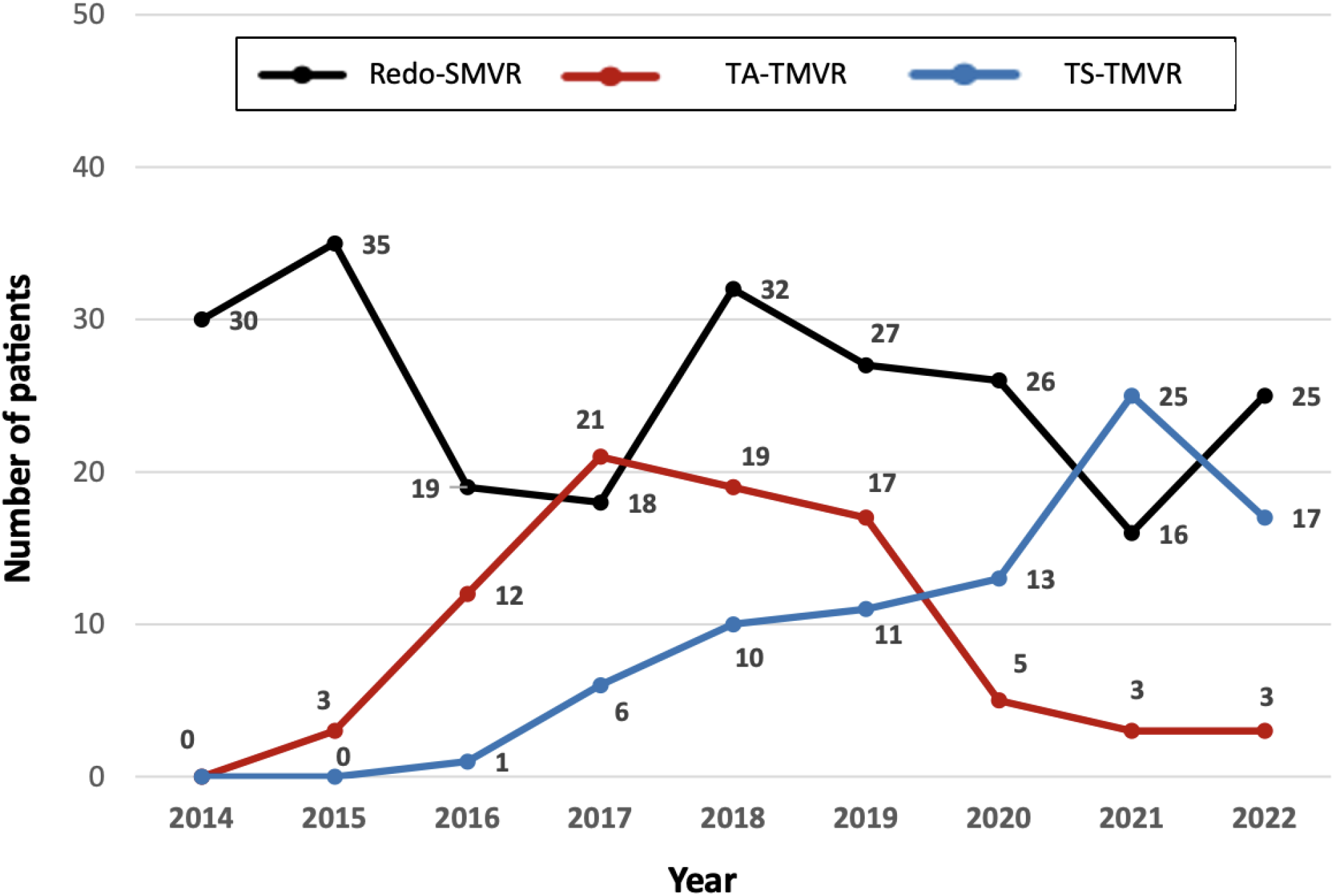
Number of procedures by groups throughout the years. Redo surgical mitral valve replacement (Redo-SMVR); Transapical Valve-in-valve transcatheter mitral valve replacement (TA-VIV-TMVR); Transseptal Valve-in-valve transcatheter mitral valve replacement (TS-VIV-TMVR)

The data were collected retrospectively for cases performed before initiation of the registry and prospectively thereafter. Formal written consents related to the procedure were obtained before each intervention. The study was approved by Institutional Review Board of the University of Sao Paulo (Heart Institute – InCor) with a waiver of patient consent.

### Doppler echocardiographic measurements

Doppler echocardiographic examination was performed before mitral intervention, upon hospital discharge, and at late follow-up. Bioprostheses dysfunction was defined according to current guidelines.^21,22^ Severe bioprosthetic stenosis was defined as a calculated mitral prosthesis area ≤ 1.0 cm^2^ or mean transmitral gradient ≥ 10 mmHg, Mitral regurgitation (MR) was defined integrating several doppler and quantitative findings.^10^ Mitral regurgitation severity was classified according to the American Society of Echocardiography guideline as: none/trace, mild, moderate, or severe.^23^

### Study devices and procedures

All transcatheter procedures were carried out using a balloon-expandable transcatheter heart valve, which was implanted through either TA or TS access, using standard techniques.^24,25^ Inovare (Braile Biomédica, São José do Rio Preto, SP, BRA) were implanted using the TA approach and Sapien XT, Sapien 3 or Sapien 3 Ultra (Edwards Lifescience, Irvine, CA, USA) for the TS approach. Inovare is a balloon-expandable valve with a chromium-cobalt stent frame with 6 sizes, ranging from 20 to 30 mm.^26^ Transcatheter heart valves (THV) were classified in three groups: small (nominal size < 25 mm), medium (25 ≤ nominal size < 28 mm) and large (≥ 28 mm), similar as used in previous studies.^27,28^ The choice of access route and valve sizes were determined based on preprocedural multidetector computed tomography (MDCT) and echocardiographic findings. Device sizing also relied on the manufacturer’s true internal diameter report and the ViV software application (B.V Valve in Valve Mitral app, http://www.ubqo.com/vivmitral).

Surgical procedures were performed via standard sternotomy, using traditional transatrial access under general anesthesia and extracorporeal circulation. Valve type was determined before the procedure, and valve size was determined intra-procedure through calibration with proprietary valve sizers. Following the procedure, patients were subsequently treated with anticoagulants or antiplatelet agents according to current clinical practice, customized to each individual patient.

### Endpoints and definitions

The primary endpoint was all-cause mortality at 30 days. The secondary endpoints included technical, device and procedural success, according to the Mitral Valve Academic Research Consortium (MVARC) criteria, and all-cause mortality at the latest follow-up.^29^ Baseline characteristics, outcomes, and procedural complications were described in accordance with MVARC consensus document.^29,30^ Echocardiographic parameters were reported according to the American Society of Echocardiography definition and MVARC document.^22,29,30^ Instead, for significant mitral stenosis (MS) we used the modified criteria, mean transmitral gradient ≥ 10 mmHg and/or an effective orifice area ≤ 1.0 cm^2^, as previously described.^27^

### Data collection

Data collection included baseline clinical and echocardiographic characteristics, procedural data and clinical follow-ups at discharge, 30-days, 12-months, and the most recent medical contact. Patient information, outcomes, and complications were gathered from the electronic medical records and our local databases. Follow-up information was obtained through a combination of medical records and telephone contacts. In instances requiring additional information, referring cardiologists, general practitioners, and patients or their families were contacted as needed. All data provided by each institution were anonymized and centrally collected. Any discrepancies and inconsistencies were promptly addressed through direct communication with local investigators.

### Statistical analysis

Patients were categorized into three groups based on their respective treatments: Redo-SMVR, TA-TMVR, and TS-TMVR. Categorical variables were reported as frequencies and examined with chi-square test or Fisher exact test. Continuous variables were tested for normality with the Kolmogorov-Smirnov test and reported as mean ± SD or median with interquartile ranges, as appropriate. Comparisons of continuous variables among the tree groups were performed with the Analysis of Variance (ANOVA) or Kruskal-Wallis test and according to distribution pattern. Similarly, Student t test or Mann-Whitney U test were used to compare continuous variables in two groups.

Bivariate analysis, including variable “treatment group” and the other covariate of interest, was performed to determine which baseline variable were predictive of all-cause mortality at 30 days. Variables with p-value ≤0.05 were included in the multivariable model. A multivariable logistic regression model with a stepwise backward selection method of significant predictors using entry and exit criterion of 0.05 and 0.10 respectively was used to identify independent predictors of all-cause mortality at 30 days.

Cumulative rates of death were calculated by using the Kaplan–Meier survival analysis and the log-rank test was used for comparison across the groups. Bivariate analysis, including variable “treatment group” and the other covariate of interest, was used to evaluate potential predictors of mid-term (1 year) mortality. A multivariable cox regression hazard model, including non-associated variables with p-value ≤0.05 in the bivariate analysis, was done to detect independent predictors of mortality at 1 year follow-up.

All tests of hypotheses were 2 sided and conducted at a 0.05 level of significance. All statistical analyses were performed using STATA/SE v.14.1. StataCorpLP, USA.

## RESULTS

### Baseline characteristics

From January 2014 and June 2023, a total of 425 consecutive patients presenting with mitral bioprosthetic valve failure were evaluated in the study. For several reasons, including concomitant treatment of aortic valve, mitral paravalvular leak as the sole cause of prosthesis dysfunction, salvage procedure, the presence of left atrial thrombus and the absence of patient data records, 10 patients were excluded. TS-TMVR was chosen for 92 patients (22.2%), TA-TMVR for 84 (20.2%) and Redo-SMVR for 239 (57.6%). Baseline characteristics are listed in **Table 1**. The groups were different in several clinical characteristics. TS-TMVR patients were older than TA-TMVR, followed by Redo-SMVR (Redo 51.7 ± 12.3, TA 64.3 ± 10.5, TS 73.7 ± 10.9 years, p<0.001) and more symptomatic (New York Heart Association [NYHA] class IV of 34.7%, 33.3%, 56.5%, respectively; p<0.001). In addition, transcatheter groups had a higher prevalence of hypertension, dyslipidemia, diabetes, chronic kidney disease (CKD), coronary artery disease (CAD) and prior pacing device (all with p<0.05). Peripheral artery disease (PAD) and pulmonary disease were more prevalent in TS-TMVR group (0.011). TA-TMVR group were more likely to have atrial fibrillation than Redo-SMVR group (p<0.001) and previous cardiac surgeries than redo-SMVR and TS-TMVR groups (p<0.001). Accordingly, in function of the higher burden of comorbidities, the median STS PROM score was higher in both TS and TA groups, with respect to the Redo-SMVR (Redo 2.9 [1.7 – 5.1], TA 6.6 [3.9 – 10.5], TS 6.5 [4.2 – 10]; p<0.001) and Euroscore II was higher in TS-TMVR group followed by TA-TMVR and Redo-SMVR groups (Redo 4.4 [3 – 6.7], TA 7.4 [4.6 – 10.7], TS 9.3 [5.4 – 17.4]; p<0.001). The mean time from a previous procedure was more than 11 years in all groups (Redo 11.5 ± 5.5, TA 12.1 ± 5.3, TS 11.5 ± 4.9 years; p=0.645).

**Table 1.**
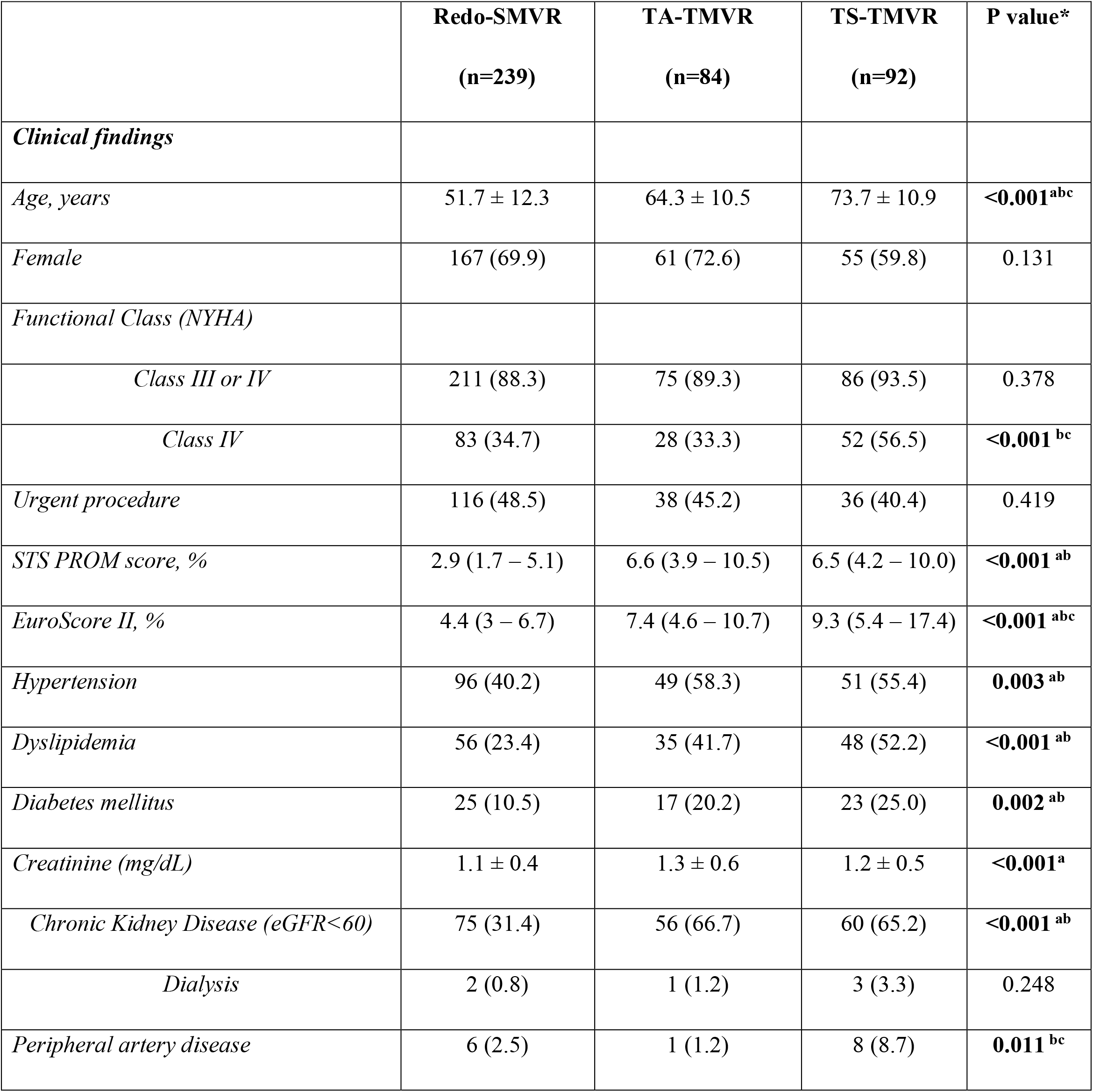
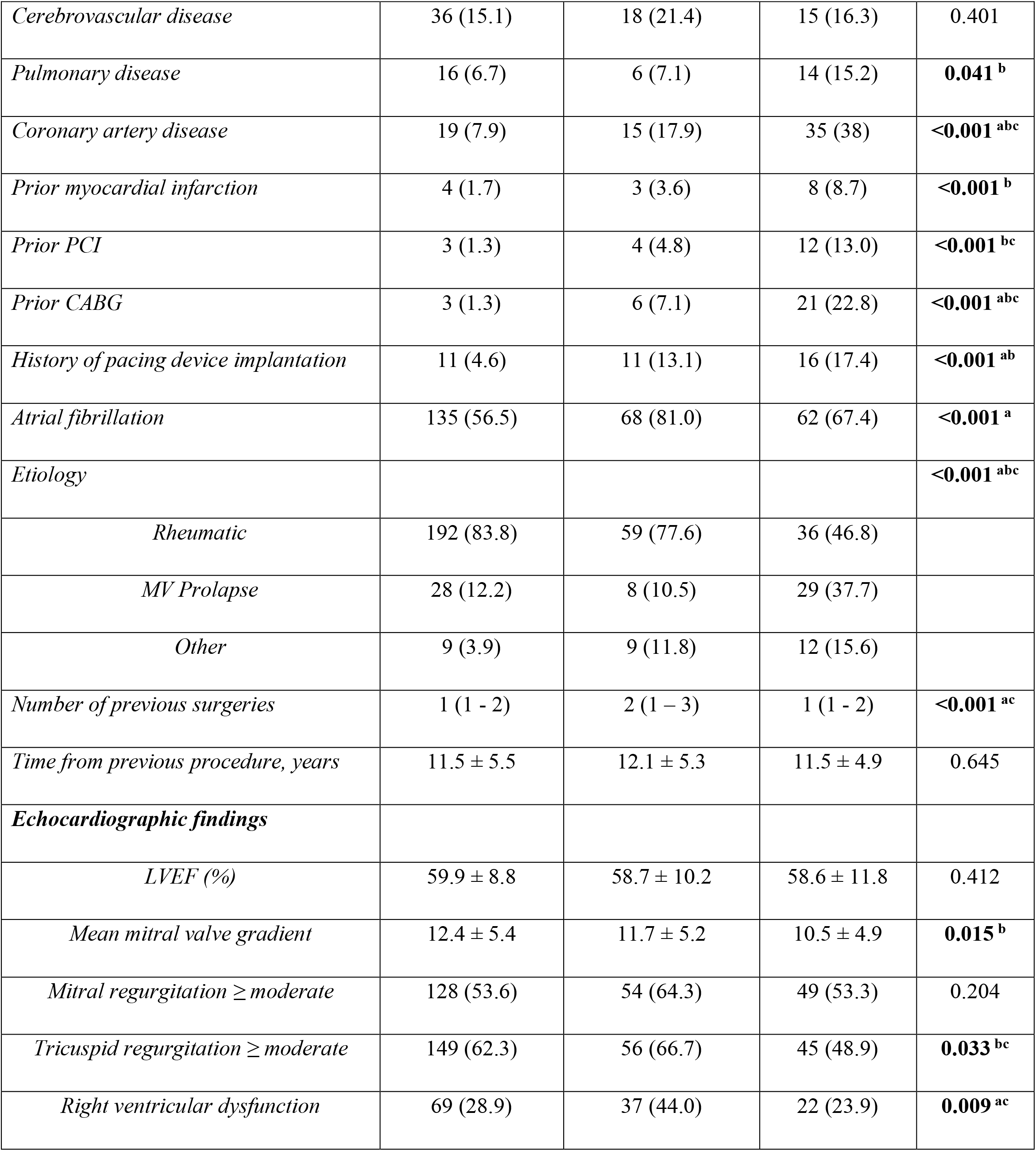
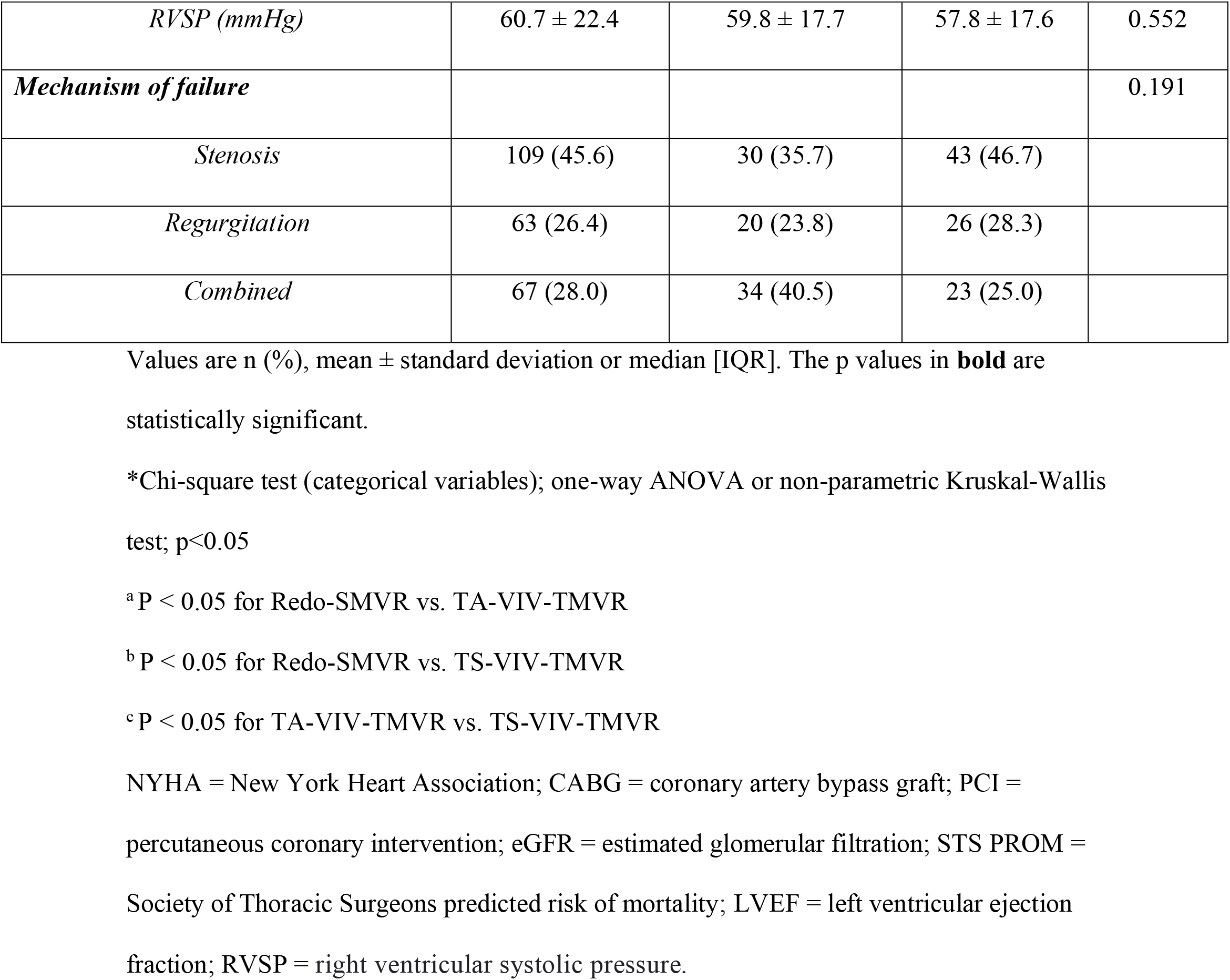
Baseline Clinical and Echocardiography Characteristics.

|  | **Redo-SMVR**<br>**(n=239)** | **TA-TMVR**<br>**(n=84)** | **TS-TMVR**<br>**(n=92)** | **P value\*** |
| --- | --- | --- | --- | --- |
| *Clinical findings* |  |  |  |  |
| *Age, years* | 51.7 ± 12.3 | 64.3 ± 10.5 | 73.7 ± 10.9 | **<0.001<sup>abc</sup>** |
| *Female* | 167 (69.9) | 61 (72.6) | 55 (59.8) | 0.131 |
| *Functional Class (NYHA)* |  |  |  |  |
| *Class III or IV* | 211 (88.3) | 75 (89.3) | 86 (93.5) | 0.378 |
| *Class IV* | 83 (34.7) | 28 (33.3) | 52 (56.5) | **<0.001<sup>bc</sup>** |
| *Urgent procedure* | 116 (48.5) | 38 (45.2) | 36 (40.4) | 0.419 |
| *STS PROM score, %* | 2.9 (1.7 – 5.1) | 6.6 (3.9 – 10.5) | 6.5 (4.2 – 10.0) | **<0.001<sup>ab</sup>** |
| *EuroScore II, %* | 4.4 (3 – 6.7) | 7.4 (4.6 – 10.7) | 9.3 (5.4 – 17.4) | **<0.001<sup>abc</sup>** |
| *Hypertension* | 96 (40.2) | 49 (58.3) | 51 (55.4) | **0.003<sup>ab</sup>** |
| *Dyslipidemia* | 56 (23.4) | 35 (41.7) | 48 (52.2) | **<0.001<sup>ab</sup>** |
| *Diabetes mellitus* | 25 (10.5) | 17 (20.2) | 23 (25.0) | **0.002<sup>ab</sup>** |
| *Creatinine (mg/dL)* | 1.1 ± 0.4 | 1.3 ± 0.6 | 1.2 ± 0.5 | **<0.001<sup>a</sup>** |
| *Chronic Kidney Disease (eGFR<60)* | 75 (31.4) | 56 (66.7) | 60 (65.2) | **<0.001<sup>ab</sup>** |
| *Dialysis* | 2 (0.8) | 1 (1.2) | 3 (3.3) | 0.248 |
| *Peripheral artery disease* | 6 (2.5) | 1 (1.2) | 8 (8.7) | **0.011<sup>bc</sup>** |
| *Cerebrovascular disease* | 36 (15.1) | 18 (21.4) | 15 (16.3) | 0.401 |
| *Pulmonary disease* | 16 (6.7) | 6 (7.1) | 14 (15.2) | **0.041<sup>b</sup>** |
| *Coronary artery disease* | 19 (7.9) | 15 (17.9) | 35 (38) | **<0.001<sup>abc</sup>** |
| *Prior myocardial infarction* | 4 (1.7) | 3 (3.6) | 8 (8.7) | **<0.001<sup>b</sup>** |
| *Prior PCI* | 3 (1.3) | 4 (4.8) | 12 (13.0) | **<0.001<sup>bc</sup>** |
| *Prior CABG* | 3 (1.3) | 6 (7.1) | 21 (22.8) | **<0.001<sup>abc</sup>** |
| *History of pacing device implantation* | 11 (4.6) | 11 (13.1) | 16 (17.4) | **<0.001<sup>ab</sup>** |
| *Atrial fibrillation* | 135 (56.5) | 68 (81.0) | 62 (67.4) | **<0.001<sup>a</sup>** |
| *Etiology* |  |  |  | **<0.001<sup>abc</sup>** |
| *Rheumatic* | 192 (83.8) | 59 (77.6) | 36 (46.8) |  |
| *MV Prolapse* | 28 (12.2) | 8 (10.5) | 29 (37.7) |  |
| *Other* | 9 (3.9) | 9 (11.8) | 12 (15.6) |  |
| *Number of previous surgeries* | 1 (1 - 2) | 2 (1 – 3) | 1 (1 - 2) | **<0.001<sup>ac</sup>** |
| *Time from previous procedure, years* | 11.5 ± 5.5 | 12.1 ± 5.3 | 11.5 ± 4.9 | 0.645 |
| ***Echocardiographic findings*** |  |  |  |  |
| *LVEF (%)* | 59.9 ± 8.8 | 58.7 ± 10.2 | 58.6 ± 11.8 | 0.412 |
| *Mean mitral valve gradient* | 12.4 ± 5.4 | 11.7 ± 5.2 | 10.5 ± 4.9 | **0.015<sup>b</sup>** |
| *Mitral regurgitation ≥ moderate* | 128 (53.6) | 54 (64.3) | 49 (53.3) | 0.204 |
| *Tricuspid regurgitation ≥ moderate* | 149 (62.3) | 56 (66.7) | 45 (48.9) | **0.033<sup>bc</sup>** |
| *Right ventricular dysfunction* | 69 (28.9) | 37 (44.0) | 22 (23.9) | **0.009<sup>ac</sup>** |
| *RVSP (mmHg)* | 60.7 ± 22.4 | 59.8 ± 17.7 | 57.8 ± 17.6 | 0.552 |
| ***Mechanism of failure*** |  |  |  | 0.191 |
| *Stenosis* | 109 (45.6) | 30 (35.7) | 43 (46.7) |  |
| *Regurgitation* | 63 (26.4) | 20 (23.8) | 26 (28.3) |  |
| *Combined* | 67 (28.0) | 34 (40.5) | 23 (25.0) |  |

Values are n (%), mean ± standard deviation or median [IQR]. The p values in **bold** are statistically significant.
\*Chi-square test (categorical variables); one-way ANOVA or non-parametric Kruskal-Wallis test; p<0.05
<sup>a</sup> P < 0.05 for Redo-SMVR vs. TA-VIV-TMVR
<sup>b</sup> P < 0.05 for Redo-SMVR vs. TS-VIV-TMVR
<sup>c</sup> P < 0.05 for TA-VIV-TMVR vs. TS-VIV-TMVR
NYHA = New York Heart Association; CABG = coronary artery bypass graft; PCI = percutaneous coronary intervention; eGFR = estimated glomerular filtration; STS PROM = Society of Thoracic Surgeons predicted risk of mortality; LVEF = left ventricular ejection fraction; RVSP = right ventricular systolic pressure.

The predominant mechanism of failure was isolated mitral stenosis (MS) among all groups (Redo 45.6%, TA 35.7%, TS 46.7%; p=0.191). Most patients had preserved ventricular function (mean left ventricular ejection fraction [LVEF] of Redo 59.9 ± 8.8%, TA 58.7 ± 10.2%, TS 58.6 ± 11.8%; p=0.412), while right ventricular (RV) dysfunction was more frequent in TA-TMVR group (p=0.009) and moderate or severe tricuspid regurgitation (TR) was more common in Redo-SMVR and TA-TMVR groups (p=0.033). RV systolic pressure (RVSP) was high but similar among the groups (Redo 60.7 ± 22.4, TA 59.8 ± 17.7, TS 57.8 ± 17.6 mmHg; p=0.552).

Rheumatic disease was the most common etiology for the initial SMVR throughout the groups, but it was even more frequent in the Redo-SMVR and TA-TMVR groups (Redo 83.2%, TA 77.6%, TS 46.8%, p<0.001). Rheumatic patients were younger and had lower prevalence of comorbidities such as dyslipidemia, diabetes, PAD, CAD and pacing devices (all with p<0.05). The median number of previous surgeries were higher in this group of patients (p<0.001), along with more moderate or severe TR (p=0.028) and any RV dysfunction (p=0.022). **(Supplementary Table 1)**

### Procedural data

Procedural data are summarized in **Table 2**. All redo-SMVR procedures were performed via median sternotomy, and 180 patients (75.3%) had the degenerated valve replaced by another bioprosthesis, the remaing patients were treated with mechanical prostheses. Surgical tricuspid valve repair was performed in 15 patients (6.3%) and left atrial appendage closure was done in 9 patients (3.8%) according to the surgeon’s judgment. During TS procedures, apical rail was not used and venoarterial loop was used in only one patient. There was no difference in terms of predilation between the groups, but there was a trend toward more post-dilation in the TS-TMVR group (p=0.062). Planned concomitant procedures during transcatheter interventions were performed in 7 patients (7.6%) that underwent mitral paravalvular leak closure and only one patient (1.1%) received a cerebral embolic protection device. THVs classified as small were implanted in 12 patients (6.8%), medium in 54 (30.7%) and large in 110 patients (62.5%).

**Table 2.**
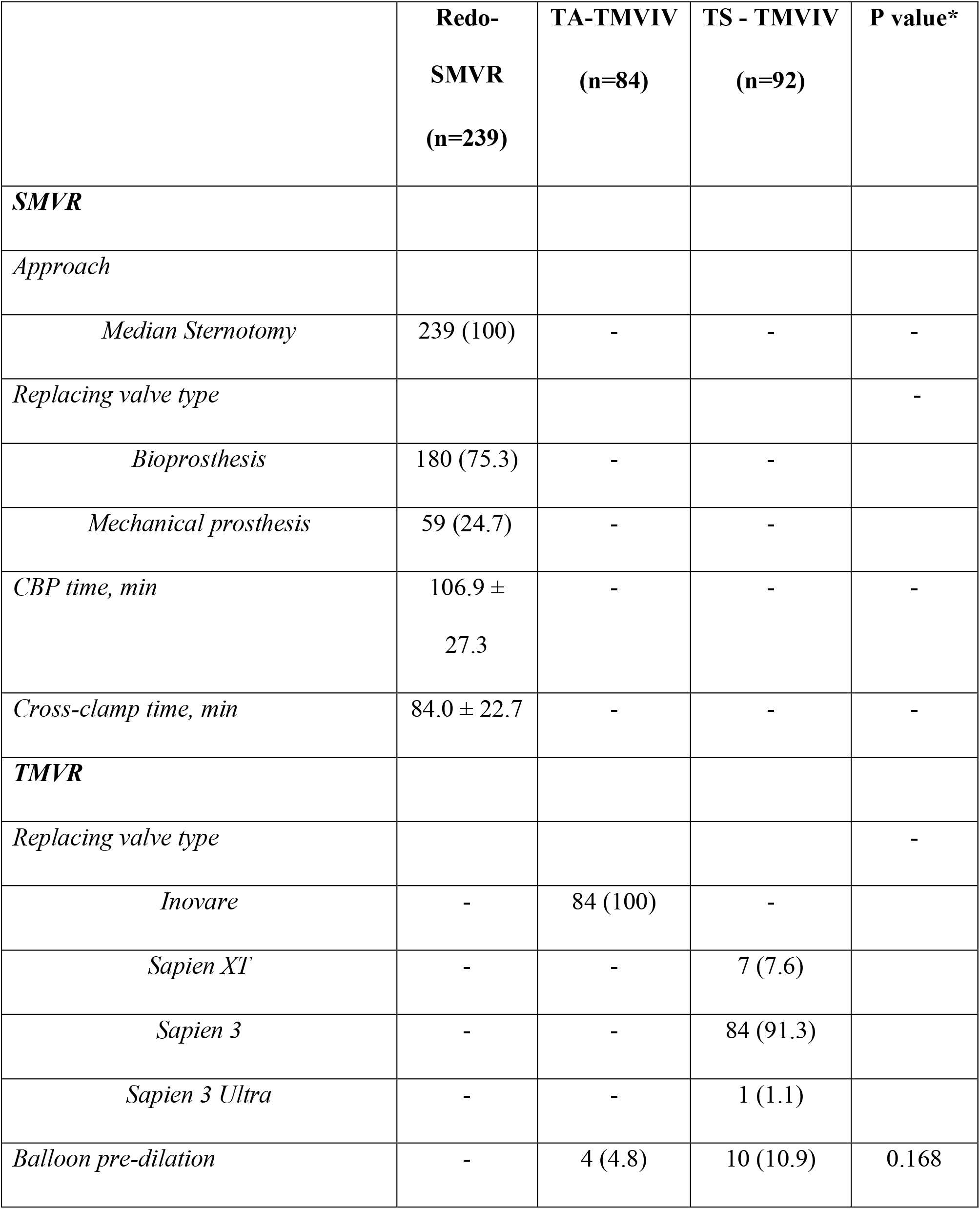
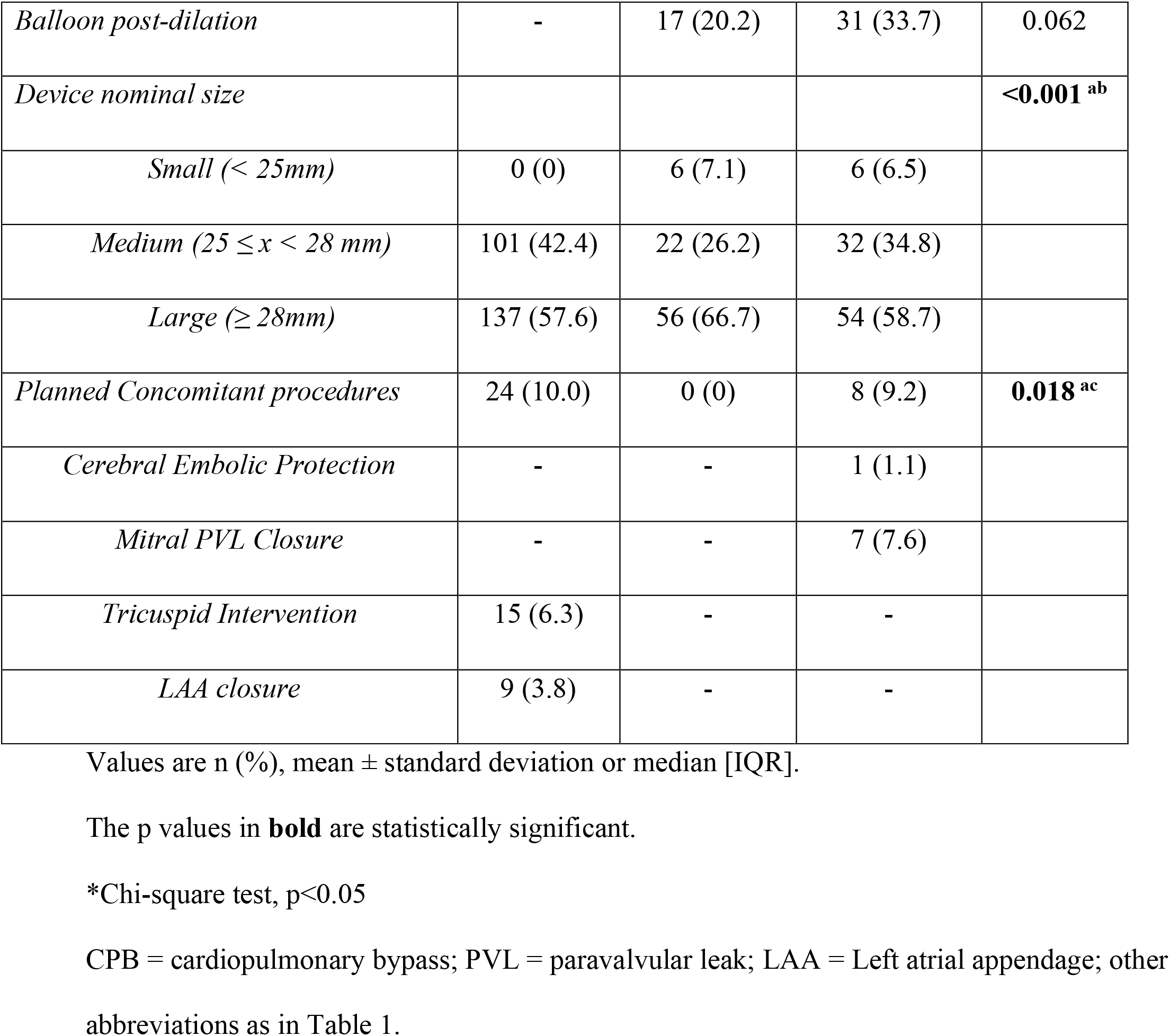
Procedural Details.

|  | **Redo-**<br>**SMVR**<br>**(n=239)** | **TA-TMVIV**<br>**(n=84)** | **TS - TMVIV**<br>**(n=92)** | **P value\*** |
| --- | --- | --- | --- | --- |
| **SMVR** |  |  |  |  |
| *Approach* |  |  |  |  |
| *Median Sternotomy* | 239 (100) | - | - | - |
| *Replacing valve type* |  |  |  | - |
| *Bioprosthesis* | 180 (75.3) | - | - |  |
| *Mechanical prosthesis* | 59 (24.7) | - | - |  |
| *CBP time, min* | 106.9 ±<br>27.3 | - | - | - |
| *Cross-clamp time, min* | 84.0 ± 22.7 | - | - | - |
| **TMVR** |  |  |  |  |
| *Replacing valve type* |  |  |  | - |
| *Inovare* | - | 84 (100) | - |  |
| *Sapien XT* | - | - | 7 (7.6) |  |
| *Sapien 3* | - | - | 84 (91.3) |  |
| *Sapien 3 Ultra* | - | - | 1 (1.1) |  |
| *Balloon pre-dilation* | - | 4 (4.8) | 10 (10.9) | 0.168 |
| *Balloon post-dilation* | - | 17 (20.2) | 31 (33.7) | 0.062 |
| *Device nominal size* |  |  |  | **<0.001**<sup>ab</sup> |
| *Small (< 25mm)* | 0 (0) | 6 (7.1) | 6 (6.5) |  |
| *Medium ($25 \leq x < 28$ mm)* | 101 (42.4) | 22 (26.2) | 32 (34.8) |  |
| *Large ($\geq 28$mm)* | 137 (57.6) | 56 (66.7) | 54 (58.7) |  |
| *Planned Concomitant procedures* | 24 (10.0) | 0 (0) | 8 (9.2) | **0.018**<sup>ac</sup> |
| *Cerebral Embolic Protection* | - | - | 1 (1.1) |  |
| *Mitral PVL Closure* | - | - | 7 (7.6) |  |
| *Tricuspid Intervention* | 15 (6.3) | - | - |  |
| *LAA closure* | 9 (3.8) | - | - |  |

Values are n (%), mean ± standard deviation or median [IQR].
The p values in **bold** are statistically significant.
\*Chi-square test, $p < 0.05$
CPB = cardiopulmonary bypass; PVL = paravalvular leak; LAA = Left atrial appendage; other abbreviations as in Table 1.

### Procedural and 30-day outcomes

Procedural and 30-day outcomes are summarized in **Table 3**. Procedural complications had rarely occurred and the MVARC technical success was high and similar among groups (Redo 98.7%, TA 96.4%, TS 96.7%, p=0.327). TA-TMVR group had one case of atrial valve embolization and one severe MR due to malpositioning requiring a second valve implantation. TS-TMVR group had two LV perforation leading to cardiac tamponade, conversion to surgery, resulting in cardiac death. Cardiac bypass (CPB) was used in all Redo-SMVR procedures and intra-aortic balloon pump (IABP) was implanted in 5.4% of Redo-SMVR group, 6% of TA-TMVR and 1.1% of TS-TMVR patients. Unplanned procedures were also rare, so that one patient from Redo-SMVR group underwent an urgent coronary artery bypass graft (CABG) procedure, three patients (3.3%) from TS-TMVR had the iatrogenic atrial septal defect closed due to hypoxemia at the end of the procedure and one patient required alcohol septal ablation due to severe LV outflow tract obstruction.

**Table 3.**
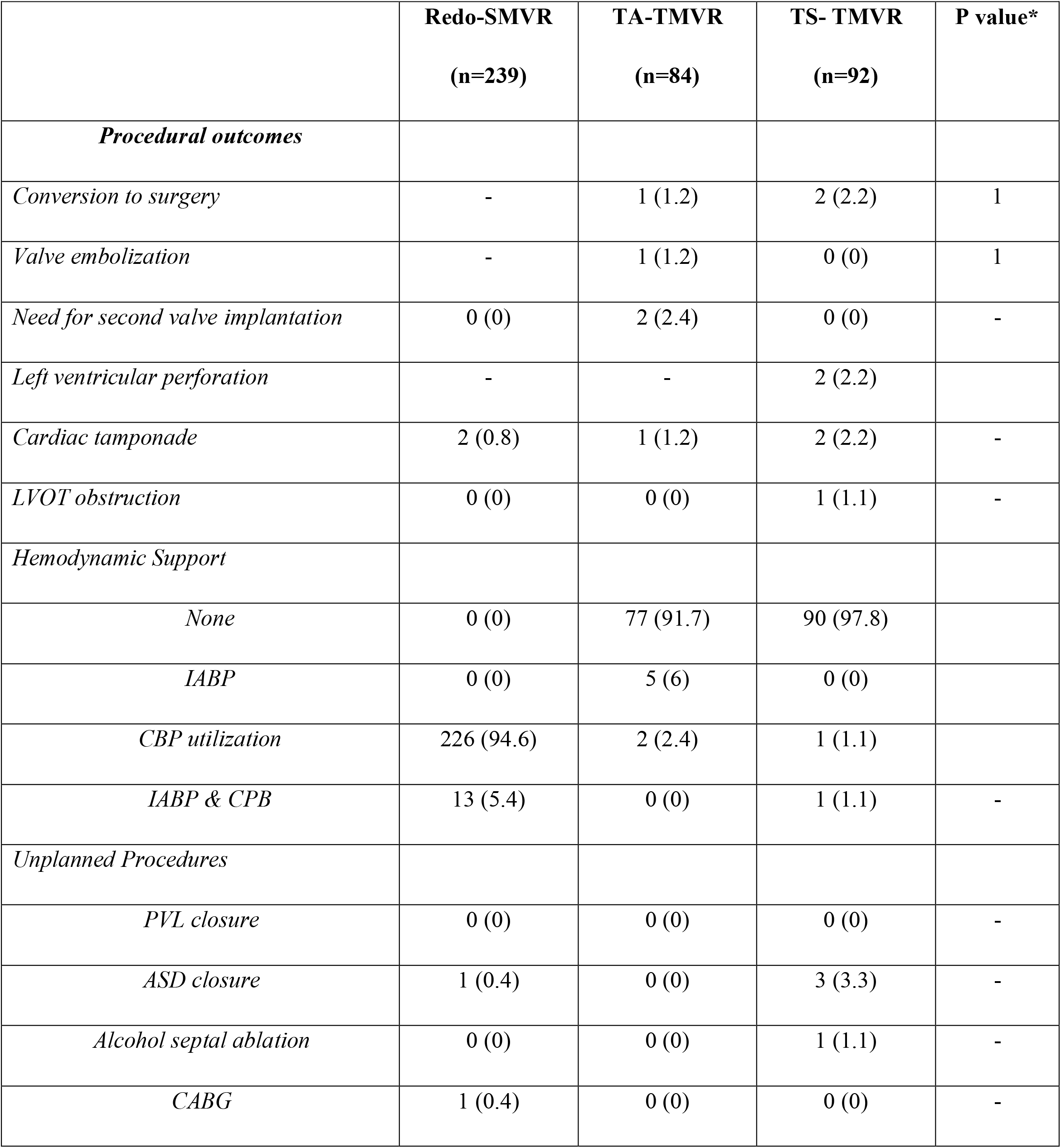
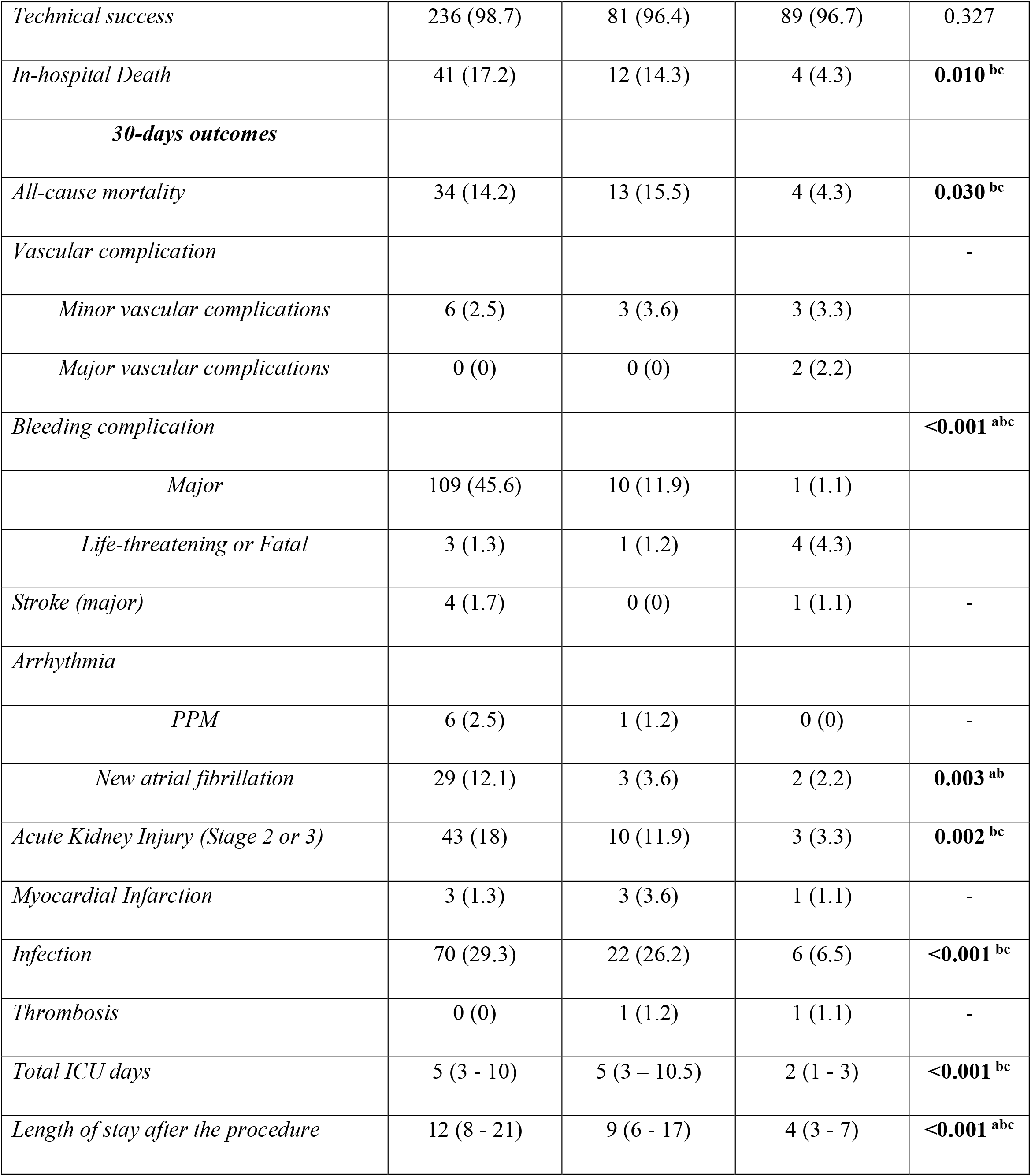
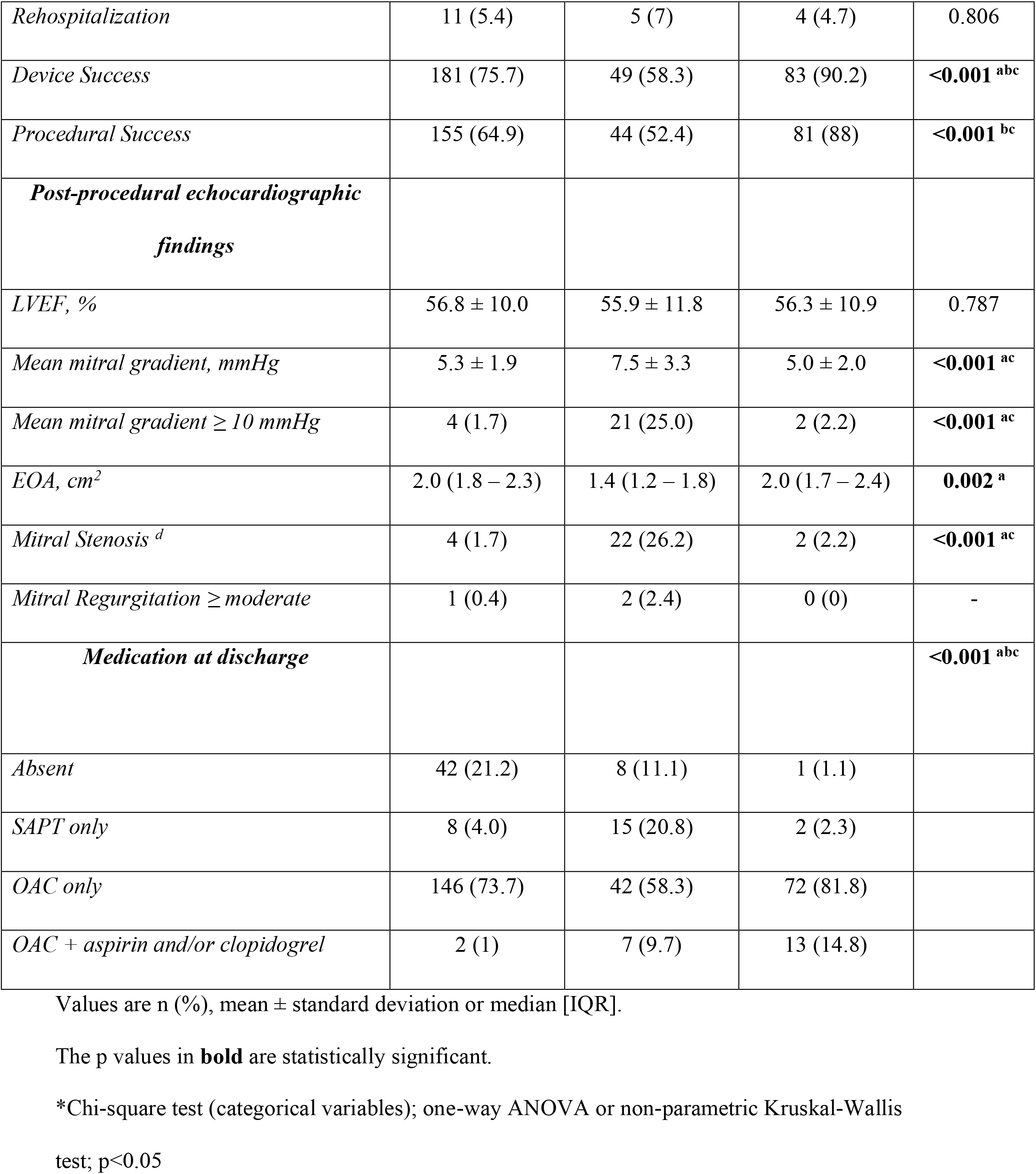
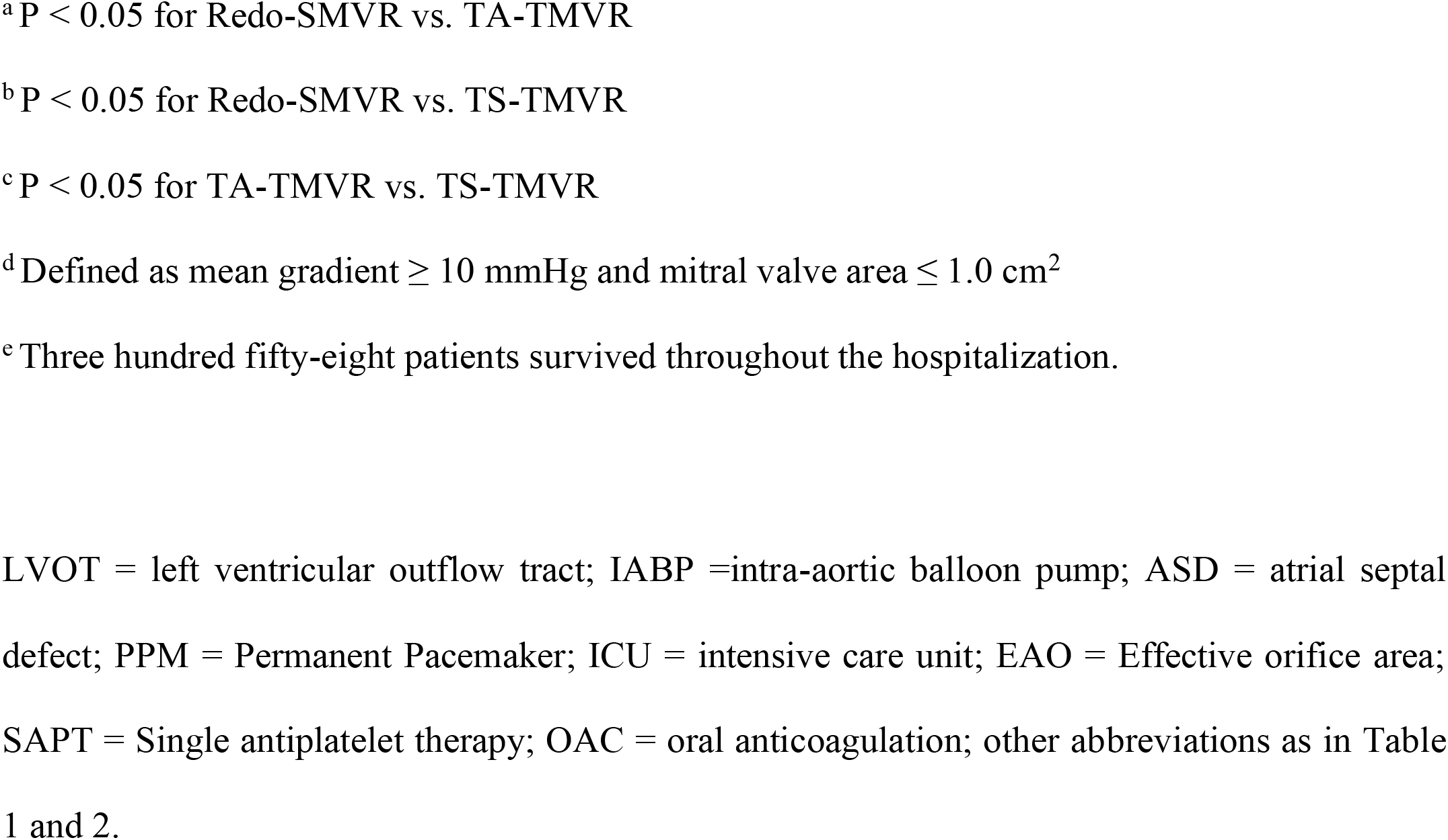
Procedural and 30-day Outcomes.

In-hospital post-procedural echocardiogram demonstrated a mean mitral valve gradient significantly higher (p<0.001) and effective orifice area (EOA) significantly lower (p<0.002) in TA-TMVR group. Mean mitral gradient greater than 10 mmHg had occurred in 25% in TA-TMVR, 2.2% in TS-TMVR and 1.7% in Redo-SMVR groups (p<0.001) and 3 patients presented with moderate perivalvular MR, one in the Redo-SMVR group and two in the TA-TMVR group.

Regarding the 30-day outcomes, severe bleeding complications were more frequent in Redo-SMVR group, followed by TA group and less frequent in TS group (p<0.001). Major vascular complications occurred in 2.5%, 3.6% and 3.3% in Redo-SMVR, TA-TMVR and TS-TMVR groups, respectively. Stroke and myocardial infarction were uncommon as well as new pacemaker implantation. New atrial fibrillation was more frequent in the Redo-SMVR group (p=0.003). The incidence of stage 2 and 3 acute kidney injury (AKI) and post-procedural infection were higher in both Redo-SMVR and TA as compared to TS groups (AKI p=0.002; post-procedural infection p<0.001).

All-cause 30-day mortality was higher in Redo-SMVR and TA-TMVR groups than in TS-TMVR (14.2% vs 15.5% vs 4.3%, respectively; p=0.030). Device success and procedural success were higher in TS-TMVR group followed by Redo-SMVR and TA-TMVR (device success: Redo 75.7% vs TA 58.3% vs TS 90.2%, with p<0.001; procedural success: 64.9% vs 52.4% vs 88%, respectively, with p<0.001). The rate of in-hospital death was also lower in TS-TMVR as compared to Redo-SMVR and TA-TMVR groups (Redo 17.2%, TA 14.3%, TS 4.3%; with p=0.010; Redo vs. TA p=0.610, Redo vs. TS p=0.002, TA vs. TS p=0.033).

As shown in Table 4, by multivariable analysis lower preprocedural LVEF, higher preprocedural RVSP and the presence of CAD were independently associated with 30-day mortality while TS-TMVR approach (using Redo-SMVR as a reference) was protective. A multivariable sensitivity analysis focusing only on patients presenting with rheumatic disease yielded similar results to the entire cohort, identifying similar predictors of short-term mortality. **(Table 5)**.

**Table 4.** Predictors of all-cause mortality.

|  | **Uni / Bivariate**<br>**Model** | **P value** | **Multivariable Model** | **P value** |
| --- | --- | --- | --- | --- |
| **30-day mortality <sup>a</sup>** | **OR (CI 95%)** |  | **OR (CI 95%)** |  |
| TA-TMVR vs Redo-SMVR (ref) | 1.10 (0.55 - 2.21) | 0.780 <sup>c</sup> | 0.81 (0.37 – 1.76) | 0.586 |
| TS-TMVR vs Redo-SMVR (ref) | 0.27 (0.09 - 0.80) | **0.017 <sup>c</sup>** | 0.12 (0.03 - 0.39) | **<0.001** |
| NYHA class IV – pre | 2.07 (1.14 – 3.79) | **0.018 <sup>d</sup>** | 1.73 (0.89 – 3.35) | 0.106 |
| CAD | 3.58 (1.68 - 7.63) | **<0.001 <sup>d</sup>** | 4.10 (1.80 – 9.34) | **<0.001** |
| LVEF (%) – pre* | 0.96 (0.93 - 0.98) | **0.001 <sup>d</sup>** | 0.95 (0.92 - 0.98) | **0.002** |
| RVSP (mmHg) – pre* | 1.02 (1.00 - 1.03) | **0.047 <sup>d</sup>** | 1.02 (1.00 - 1.03) | **0.019** |
| **1-year mortality <sup>b</sup>** | **HR (CI 95%)** |  | **HR (CI 95%)** |  |
| TA-TMVR vs Redo-SMVR (ref) | 1.01 (0.57 - 1.80) | 0.965 <sup>c</sup> | 0.80 (0.45 - 1.44) | 0.455 |
| TS-TMVR vs Redo-SMVR (ref) | 0.56 (0.28 - 1.11) | 0.098 <sup>c</sup> | 0.38 (0.18 - 0.80) | **0.011** |
| LVEF (%) – pre* | 0.96 (0.95 - 0.98) | **<0.001 <sup>d</sup>** | 0.96 (0.94 - 0.98) | **<0.001** |
| TR ≥ moderate – pre | 2.09 (1.20 - 3.62) | **0.009 <sup>d</sup>** | 2.36 (1.35 - 4.12) | **0.003** |
| CAD | 2.36 (1.30 - 4.26) | **0.005 <sup>d</sup>** | 2.66 (1.46 - 4.86) | **0.001** |

<sup>a</sup> Logistic regression and Wald test, p<0.05
<sup>b</sup> Cox regression model and Wald test, p<0.05.
<sup>c</sup> Univariate analysis.
<sup>d</sup> Bivariate analysis.
\*With each additional unit
Abbreviations as in Tables 1, 2 and 3.

**Table 5.** Multivariable sensitive analysis considering only rheumatic patients for 30-day and 1-year mortality.

| **30-day mortality <sup>a</sup>** | **Multivariable Model** | **OR (CI 95%)** | **p\*** |
| --- | --- | --- | --- |
| Treatment group | TA-TMVR vs Redo-SMVR (ref) | 0.44 (0.15 - 1.33) | 0.148 |
|  | TS-TMVR vs Redo-SMVR (ref) | 0.07 (0.01 - 0.58) | **0.014** |
| NYHA class IV – pre | Yes x no (ref) | 2.26 (0.98 - 5.23) | 0.056 |
| CAD | Yes x no (ref) | 7.71 (2.53 - 23.5) | **<0.001** |
| LVEF (%) – pre | With each additional unit | 0.94 (0.90 - 0.97) | **0.001** |
| RVSP (mmHg) – pre | With each additional unit | 1.02 (1.00 - 1.04) | **0.016** |
| **1-year mortality <sup>b</sup>** | **Multivariable Model** | **HR (CI 95%)** | **p\*** |
| Treatment group | TA-TMVR vs Redo-SMVR (ref) | 0.57 (0.26 - 1.26) | 0.162 |
|  | TS-TMVR vs Redo-SMVR (ref) | 0.28 (0.08 - 0.95) | **0.041** |
| LVEF (%) – pre | With each additional unit | 0.96 (0.94 - 0.99) | **0.005** |
| TR ≥ moderate - pre | Yes x no (ref) | 2.65 (1.23 - 5.71) | **0.013** |
| Coronary artery disease | Yes x no (ref) | 3.64 (1.68 - 7.92) | **0.001** |

<sup>a</sup> Logistic regression and Wald test, p<0.05
<sup>b</sup> Cox regression model and Wald test, p<0.05.

Echocardiographic data at early follow-up, up to 30 days, **is described in Supplemental Table 2**. Although there were significant differences in antithrombotic therapy at discharge across the 3 groups, oral anticoagulation alone or in combination with antiplatelet agents was the preferred regimen for most patients, irrespective of the treatment group. In Redo-SMVR group, 21.2% of patients did not receive any antithrombotic therapy and only one patient was treated with a combination of oral anticoagulation and antiplatelet therapy. Oral anticoagulation was prescribed to 68% of patients in the TA-TMVR group, which represented the lowest rate of anticoagulation when compared to the other groups (Redo 74,7% and TS 96.6%, p<0.001). (**Table 3**) Prosthesis thrombosis was an infrequent complication, with only two occurrences within 30-day follow-up: one in TS-TMVR and one in TA-TMVR groups. Both patients were successfully managed with anticoagulation therapy.

### 1-year follow-up and predictors of mortality

Over a median follow-up period of 489 days (interquartile range 86 – 1074 days), a total of 85 patients had died, 46 being patients in the Redo group, 19 patients in TA-TMVR group and 20 in the TS-TMVR group. There was no difference in terms of 1-year mortality rate across the groups (19.8% vs. 20.0% vs 12.5%, respectively, log-rank 0.222; **Figure 2A**). A landmark analysis demonstrated lower rate of all-cause mortality at 30 days (14.4% vs. 15.7% vs 4.4% in Redo, respectively, log-rank 0.035) and similar mid-term mortality (30–360 days) across the groups (5.6% vs 5.1% vs 8.6%, respectively, log-rank 0.649; **Figure 2B**). Cardiac mortality responded for 95,3%, 75% and 70% in Redo, TA-TMVR and TS-TMVR groups respectively.

**Figure 2.**
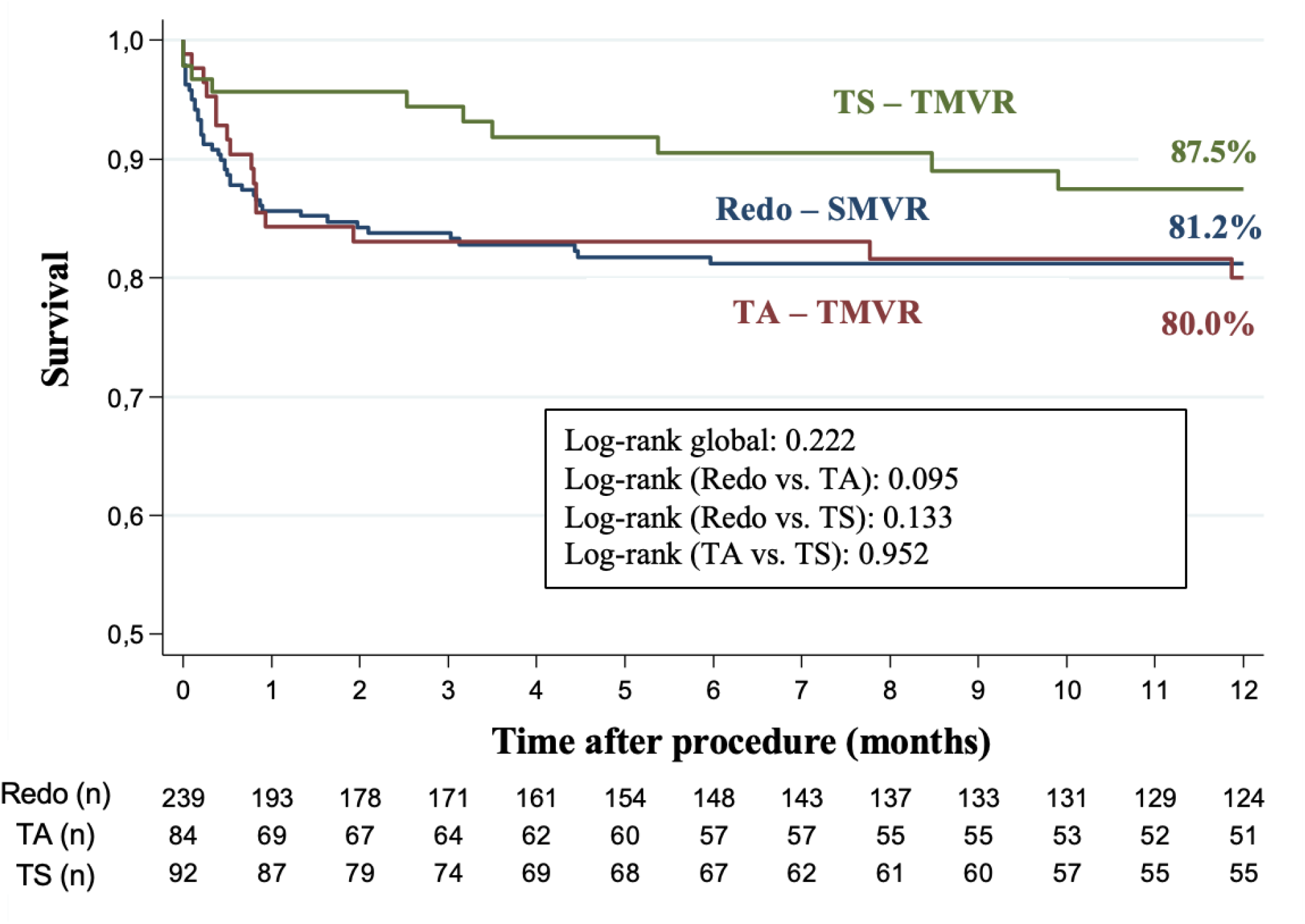
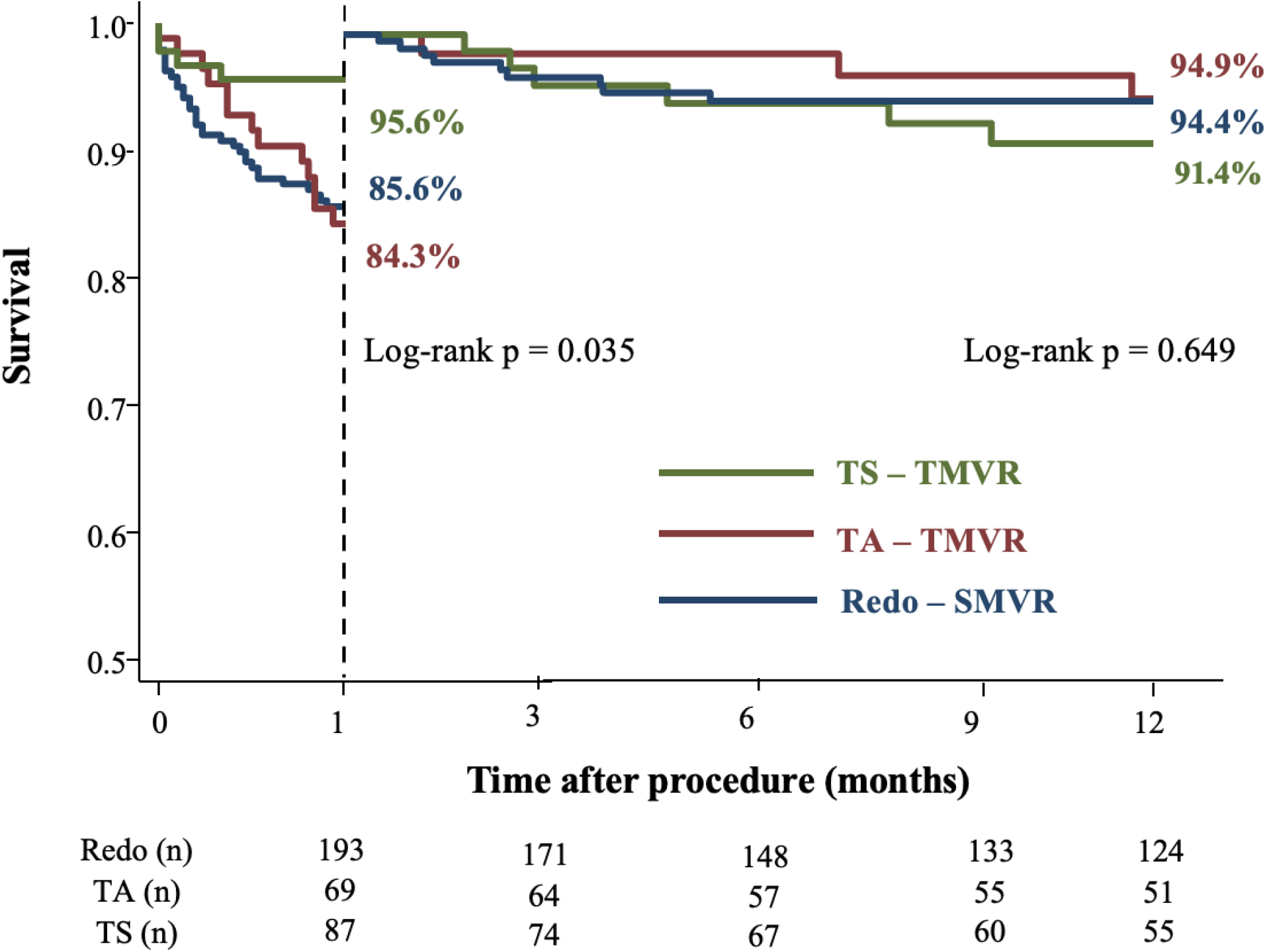
Survival analysis according to the treatment group. (A) The survival rate of patients undergoing TS-VIV-TMVR (green line), Redo-SMVR (blue line), and TA-VIV-TMVR (red line) are shown. (B) The survival rate with landmark analyses (0–30 days and 30–360 days) showed reduced early mortality (0–30 days) in TS-VIV-TMVR group but no difference was found in late mortality (30–360 days) after Redo-SMVR, TS-VIV-TMVR and TA-VIV-TMVR.

**Table 4** shows the multivariable analysis for 1-year all-cause mortality. In this analysis, lower preprocedural LVEF, the presence of CAD and the presence of significant TR (moderate or severe) remained independently associated with mid-term mortality while TS-TMVR approach was protective. Following a sensitivity analysis focused on patients with rheumatic disease, the predictors of 1-year all-cause mortality remained consistent with those observed in the total population. **(Table 5)**

The 1-year survival analysis based on the post-procedural mean mitral gradient of patients discharged from the hospital after the procedure, divided into three strata (<5; 5 – 9,9; >=10 mmHg), revealed no differences in terms of mortality across the three ranges of gradients. Considering the potential relevance of this variable, a similar analysis was conducted using it as continuous data and dividing the population into two groups using different cut-offs. No differences were found in any of the described analyses. (Supplementary Figure 1)

Late echocardiographic data were available from 132 patients, 60 in Redo-SMVR group, 35 in TA-TMVR and 37 in TS-TMVR with a median time from index procedure of 778 (537-1173) in Redo-SMVR, 719 (469-929) in TA-TMVR and 398 (374-615) days in TS-TMVR groups. **(Supplemental Table 2**)

Until the latest follow-up (after 30-day), we detected three additional prosthesis thrombosis associated with worsening symptoms, two in TS group and one of than in TA group. One patient was on single antiplatelet therapy (SAPT), another on direct oral anticoagulation with SAPT and the third was on warfarin alone. These patients were treated with adjustments in anti-thrombotic therapy without surgical intervention. Considering the whole follow-up period, the occurrence of clinical prosthesis thrombosis was higher in patients treated with transcatheter procedures compared to the Redo-SMVR group (TMVR 2.9%, Redo-SMVR 0%; p=0.030).

## DISCUSSION

The major findings of the present study are as follows: (i) The TS-TMVR group presented lower rates of periprocedural complications and 30-day mortality compared to both the Redo-SMVR and TA-TMVR groups, as well as higher MVARC procedural success, despite being older and with a higher burden of comorbidities and predicted surgical risk; (ii) lower LVEF, higher RVSP and the presence of CAD were independent predictors of 30-day all-cause mortality while TS approach was protective; (iii) lower LVEF, presence of CAD and presence of significant TR (moderate or severe) were independently associated with 1-year mortality, while TS-TMVR treatment was protective; (iv) Patients with rheumatic disease had similar predictors of short-term and mid-term mortality as compared to those observed in the entire population.

Redo-SMVR is still the treatment of choice for patients with dysfunctional mitral bioprosthesis, yet the risk of a repeated mitral valve surgery is high, and it increases in each additional redo-SMVR as demonstrated by previous studies.^31^ Therefore, TMVR using the TA and TS approaches has emerged as an alternative in such patients, with prior data showing its safety and efficacy in moderate and high-risk patients. ^32-34^ Our study extends further such results, with larger number of patients and events, and with a direct comparison of the transcatheter approach used (TA vs. TS), against the surgical standard (Redo-SMVR). Of note, TS-TMVR was related to a higher MVARC procedural success and better short-term results, including a 3-fold lower 30-day mortality, despite a much higher risk profile. Using the Redo-SMVR group as a reference, TS strategy was identified as an independent predictor of lower 30-day mortality by multivariable analysis. Additionally, the presence of CAD, lower preprocedural LVEF and higher RVSP were independently associated with all-cause mortality at 30-days.

As described in previous studies, all-cause mortality at 30-day varied in a very wide range from 3.4 to 17.2% in Redo-SMVR patients, from 4.5 to 11.9% in the TA and from 3.3 to 11.1% in TS approach. ^11,19,20,32,33,35^ Likewise, these results have been gathered in several recent meta-analyses, and despite one of them has demonstrated a lower incidence of in-hospital mortality with the transcatheter strategies (3.2% vs. 6.8%, p<0.001), until now there was no significant difference between Redo-SMVR versus transcatheter treatment in terms of 30-day all-cause mortality. ^35,36^ Additionally, the TS technique showed a 2-fold lower median intensive care unit and in-hospital length of stay after the procedure.

Procedural complications, including valve embolization, left ventricular perforation and left ventricular outflow tract (LVOT) obstruction, were infrequent, despite that many of the included centres were in their initial experience with TMVR, each complication occurred in less than 2.5% of patients. The lower incidence of complications yielded higher incidence of MVARC technical success, what is consistent with previous studies.^12,20,33^ In addition, at 30-days, significant differences with regard to complications were observed, including bleeding, atrial fibrillation and acute kidney injury that were more frequent in Redo-SMVR than in both transcatheter groups. Importantly, such high rates of severe bleeding complications in the Redo-SMVR group (45.6%) were similar to the incidence described by Kamioka et al.^15^ (45.8%) and lower than described by Zogg et al.^19^ (61.9%). Infections occurred more frequently in both the Redo-SMVR and TA-TMVR groups than in the TS group, a finding that coincides with the incidence published by Zahid et al.^18^ likely related to the thoracotomy is such patients. New atrial fibrillation and AKI stage 2 or 3 (MVARC definition) were also significantly more frequent in the Redo-SMVR group.

The differences between the baseline characteristics across the groups are commonly seen in registries that compare surgical treatment to transcatheter approach.^15,18-20,37^ The high prevalence of rheumatic disease in this study could be an additional factor that contributed to a lower mean age, especially in Redo-SMVR and TA-TMVR groups. To our knowledge, this is the largest cohort of rheumatic patients undergoing TMVR to date. In our study, rheumatic disease was not related to early and mid-term mortality, and this population presented the same predictors of 30-day mortality as the whole cohort.

In the mid-term follow-up (∼1-year), despite being numerically lower in the TS-TMVR group, there were no significant differences in all-cause mortality across the groups, similar to the results of a recent meta-analyses with rates between 12.5 and 20%. ^35,36^ In the multivariable analysis for 1-year mortality, TS treatment remained an independent protective strategy, similarly as demonstrated in the 30-day multivariable analysis. Also, clinical features such as preprocedural LVEF and the presence of CAD continued to be independent predictors of 1-year mortality, along with moderate or severe TR. In general, in rheumatic patients, the predictors of 1-year mortality were the same as in the global population. Considering the improved mortality outcomes in both short- and mid-term analysis with TS approach vs. TA-TMVR and Redo-SMVR, one might argue that the TS-TMVR should be preferentially selected among patients with dysfunctional mitral bioprosthesis at higher risk, such as those with reduced LVEF, CAD and reduced RV functional status, such as severe TR or high RVSP. A potential advantage of the Redo-SMVR over the transcatheter strategies is the possibility to treat the tricuspid regurgitation during the same procedure (62.3% of moderate or severe TR in the Redo-SMVR), although only the minority (6.3%) underwent concomitant tricuspid intervention during the index procedure. While it is possible that this fact could potentially hinder the mid-term benefits of Redo-SMVR over the transcatheter strategy, the low rate of tricuspid intervention in this study aligns with rates reported previously by Szlapka et al. (5.7%) and Kamioka et al. (13.6%). ^15,38^

The post-procedure mean mitral gradient observed in our study was consistent with findings from previous studies, with a mean gradient frequently exceeding 5 mmHg across all groups. ^15,20,39^ Importantly, Simonato et al. reported in 2021 that there was no difference in mid-term survival when comparing two groups of patients using a 10-mmHg cut-off.^10^ Similarly, despite the smaller population size in our study, we did not observe any difference when using two different cut-offs (5 mmHg and 10 mmHg) or when dividing the population into three groups, as shown in Supplementary Figure 1. The confirmation that residual gradients did not impact mortality is very important, but it does not mean that it is not related to heart failure symptoms improvement, hospitalization or quality of life, what need further investigation.

Comparing these findings across the treatment groups, in our study the mean gradient of the TA-TMVR was greater than Redo-SMVR and TS-TVMR groups and the effective orifice area was lower in transcatheter groups. In the TA-TMVR group, 25% of patients had the mitral mean gradient greater than 10 mmHg versus 1.7% in Redo-SMVR and 2.2% in TS-TMVR group. One of the possible reasons for this difference is that, in this series, we included the very early experience with these types of procedures, and the TMVR technique has evolved over the years. ^24,40-42^ The TA approach was initiated earlier in this series, which could have led to fewer post-dilation and valve fractures during the procedure, culminating in this worse performance. It’s important to note that the transcatheter groups were treated with different prostheses and more data are necessary to determine whether the higher gradients among TA procedures are related to the implantation technique or to certain THV characteristics of the Inovare bioprosthesis as compared to the Edwards Sapien family utilized in the transseptal approach. This higher gradient in the transapical group was observed across all the spectrum of THV sizes.

## Study limitations

This study had the inherent limitation of being an observational study, with adverse events not adjudicated by an independent committee, and there is a potential for underreporting. Data for the Redo-SMVR and TA groups were collected from a single reference center in cardiovascular disease, while data for the TS group derived from multiple centers. As a non-randomized study, our findings are subject to potential selection bias and confounding factors. The large differences in baseline characteristics are impossible to correct for and the numbers are too small for propensity score matching.

The therapeutic strategy for each patient was based on the local’s Heart Team decision, potentially introducing selection bias. Echocardiographic findings were not assessed by an independent echocardiographic core laboratory.

We included patients from the earliest experiences with this type of procedure, resulting in an asymmetrical period of inclusion at the beginning. Additionally, technical improvements occurred over these years, meaning the outcomes for patients treated during this phase may not reflect contemporary results with these techniques. The transcatheter groups had implanted two different brands of balloon-expandable prostheses, while Redo-SMVR patients received either mechanical or biological prostheses.

## CONCLUSIONS

TS transcatheter mitral valve-in-valve replacement was associated with better short-term outcomes, including 30-day all-cause mortality and MVARC procedural success, compared to Redo-SMVR and the TA transcatheter approach, despite the higher-risk profile of this patients. At 1-year, there was no significant difference in mortality across the groups. Lower LVEF and the presence of CAD or significant TR were independent predictors of 1-year mortality while TS-TMVR strategy was protective. Therefore, TS-TMVR should be considered the safest strategy for treating higher risk patients with failed mitral bioprostheses, including impaired LVEF and older age, among other factors. Rheumatic patients, that were the majority in the present analysis, shared similar findings as compared to the entire cohort, so that the etiology of the mitral valve disease was rather not related to the clinical outcomes. Randomized trials and longer-term follow-up are still necessary to determine the best lifetime management strategies for this challenging population.

## Data Availability

All data generated or analyzed during this study are included in this published article and its supplementary information files. Additional data are available from the corresponding author upon reasonable request

## Abbreviations and acronyms

LVEF: Left ventricular ejection fraction
MVARC: Mitral Valve Academic Research Consortium
NYHA: New York Heart Association
TA: Transapical
TMVR: Transcatheter mitral valve replacement
TS: Transseptal
SMVR: Surgical mitral valve replacement
STS PROM: Society of Thoracic Surgeons Predicted Risk of Mortality

## Central Illustration

Transseptal transcatheter valve-in-valve implantation versus transapical and redo surgical replacement of degenerated mitral bioprosthesis.

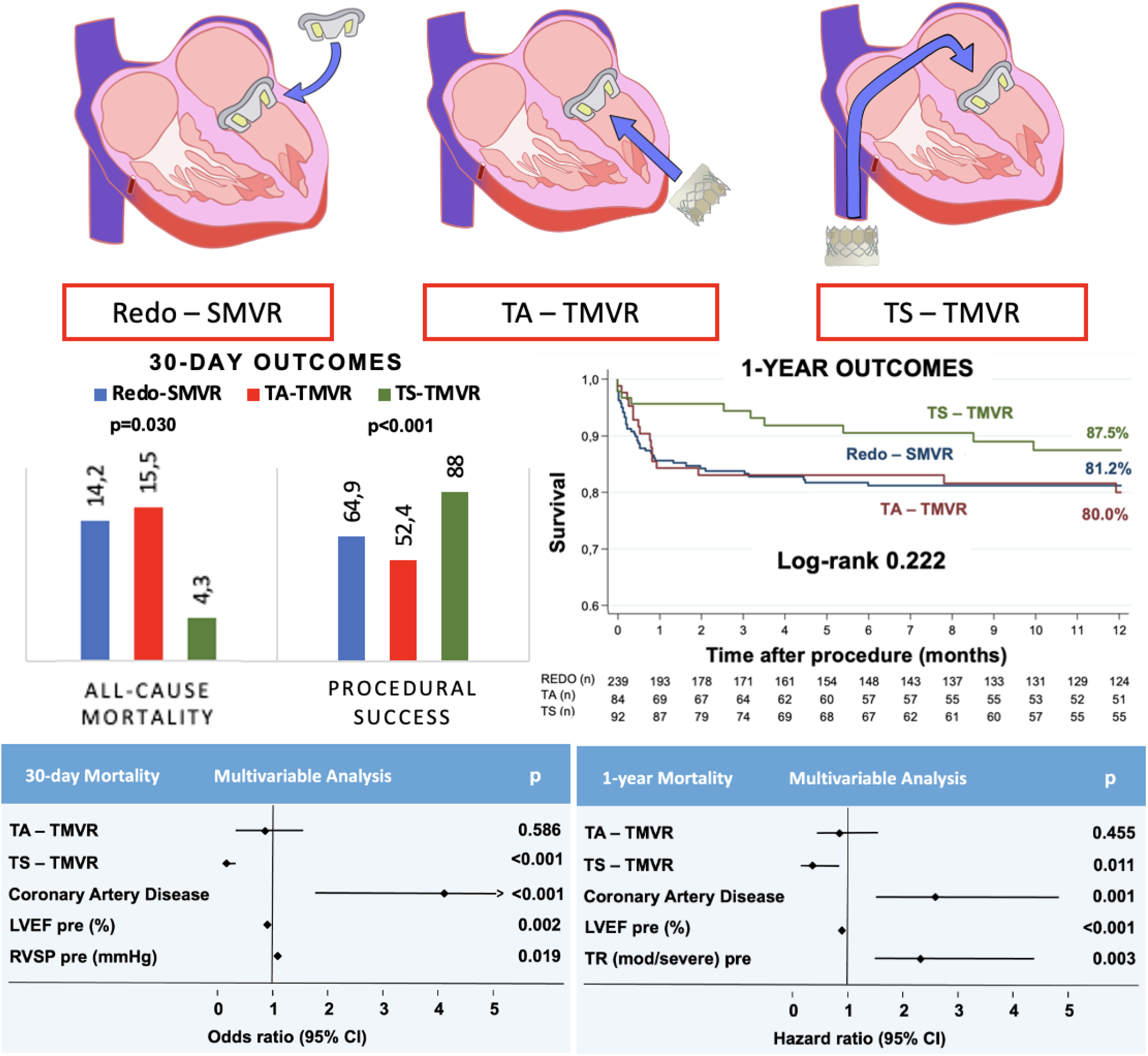

## Disclosures

Drs. Nicz, Ribeiro, Arruda, Cristovao, BAA Falcao, Martins, Sarmento-Leite, Bezerra and de Brito Jr. have served as a proctor for Edwards implantation.

Dr. Fonseca have served as a proctor for Braile Biomedica.

All other authors have reported that they have no relationships relevant to the contents of this paper to disclose.

## Notes

### Funding Statement

no external funding was received

### Author Declarations

This study was approved by the USP - HOSPITAL DAS CLÍNICAS DA FACULDADE DE MEDICINA DA UNIVERSIDADE DE SÃO PAULO - HCFMUSP Ethics Committee, under protocol number CAAE: 42627121.7.0000.0068, on March 10th, 2021.

